# Remote measurement technologies for depression in young people: A realist review with meaningful lived experience involvement and recommendations for future research and practice

**DOI:** 10.1101/2022.06.16.22276510

**Authors:** Annabel E L Walsh, Georgia Naughton, Thomas Sharpe, Zuzanna Zajkowska, Mantas Malys, Alastair van Heerden, Valeria Mondelli

## Abstract

**Background:** Remote measurement technologies (RMT), such as smartphones and wearables, allow data collection from an individual in real-time during their day-to-day life, from which their mood, physiology, behaviour, and environment can be inferred. As such, RMT could monitor and detect changes relevant to depression for objective screening, symptom management, relapse-prevention, and personalised interventions. Whilst RMT for depression in young people has been previously reviewed, technological capability and digital mental health literature steeply increase each year but with limited scrutiny of the realist and ethical considerations likely to impact the benefits, implementation, and overall potential of RMT in the real-world.

**Methods:** A realist review of RMT for depression in young people aged 14 – 24 years was conducted in collaboration with two young, lived experience co-researchers from The McPin Foundation Young People’s Network (YPN) and in accordance with the Realist and Meta-narrative Evidence Syntheses: Evolving Standards (RAMESES) for quality and publication. Iterative searches across 10 electronic databases and 7 sources of grey literature, fine-tuning of selection-criteria, data extraction and evidence synthesis with insights from the wider YPN members allowed gradual refinement of an initial framework into a realist intervention theory.

**Results:** Of 6118 records identified, 104 were included in evidence synthesis. *What does and does not work?* Smartphones were most preferred, with both passive and active data collection for a holistic approach but a balance between data quality, intrusiveness, and data privacy. From the evidence currently available, depression was best detected by changes in sleep, mobility, smartphone use, social communication, and self- or- parent-reported mood. This had some uses in screening, self-monitoring, and feedback to the healthcare professional but not in relapse-prevention and personalised interventions, where significantly more research is required. *How and why?* The impact of RMT as an intervention itself on depression outcomes remained unclear but self-monitoring and feedback improved emotional self-awareness, therapeutic relationship, and help-seeking behaviours. *For whom?* With limited standardisation and investigation of the impact of depression on adherence rates, there may be an overestimation of how much young people are likely to use RMT in the real-world. However, they were most likely to benefit those interested in and motivated by the data-driven nature, who have lower depression severity, no co-morbidities where self-monitoring could cause harm, and the presence of changeable behaviours. *In what contexts?* RMT facilitated monitoring during transition to university, known to be associated with worsening depression in young people; however, there were significant challenges in health care and school settings. Adaptability was important, such that RMT were culturally compelling and accurate for the local context. Overall, there were many gaps in the evidence and common methodological issues across the literature.

**Conclusions:** From the evidence base and lived experience insights, realist and ethical considerations were highlighted, as well as the remaining gaps in evidence and methodological issues common across the literature. For RMT to be the scalable solution for depression in young people rather than a case of overplayed potential, several important recommendations for future research and practice were made.

## Introduction

The use of digital technology is becoming increasingly common in mental health. Remote measurement technologies (RMT), such as smartphones and wearables, have been suggested as a solution to improve the monitoring, detection, and treatment of depression in young people across the globe. However, the benefits, implementation, and overall potential of RMT in the context of realist and ethical considerations remain unclear.

### The Burden of Depression in Young People & Low-resource Settings

Depression is now a leading contributor to the global burden of disease with significant impacts for the individual, society, and economy. It often begins in adolescence or young adulthood ^1,2^ reflecting a developmentally sensitive period and has a chronic course with recurrent depressive episodes throughout one’s lifetime. However, diagnosis can be challenging with a lack of objective, reliable diagnostic criteria, and reliance on retrospective self-report influenced by both recall and mood-state associated biases ^3^. Young people are particularly vulnerable to deterioration due to barriers in help-seeking, including delay in or lack of access to services, stigma, and difficulty identifying or expressing concerns ^4^, often exacerbated by poor communication, disregard of agency ^5^, and mismanagement ^6,7^. As such, many cases go undetected and untreated, contributing to a trajectory towards full-blown, chronic depression associated with poorer outcomes in adulthood ^8^. Indeed, it has been estimated that only 28% of all individuals with depression receive treatment, which decreases to just 7-14% in low- and middle-income countries (LMICs) ^9^ where almost 90% of the global adolescent population reside ^10^. For those that do receive treatment, clinical improvement is modest at best ^11,12^ with little understanding of the active ingredients of effective interventions and why they may work for some but not others ^13–15^. There is a crucial need for effective early interventions targeting the adolescent period, that can be personalised and are scalable across different contexts if we are to improve depression outcomes across the globe ^16^. Remote measurement technologies (RMT) may represent an increasingly ubiquitous and accessible resource for such an intervention.

### Remote Measurement Technologies

RMT include any digital technology with the capability to collect data from an individual in real-time during their day-to-day life through a remote interface, e.g., smartphones, wearables, and associated apps. Data collection can be active via direct input by the individual (e.g., ecological momentary assessment (EMA), mood logs) and / or passive via embedded sensors or during interaction with the device (e.g., heart rate, heart rate variation, skin conductance, actigraphy, accelerometery, global positioning system (GPS), ambient light, microphone, and paradata - screen status, app usage, call / SMS logs). Extracted data features are then used to infer the mood, physiology, behaviour, and environment of the individual (e.g., stressful events and responses, rest/activity cycles, sleep, mobility, physical and social activity, speech patterns) ^17,18^, which may also be indicative of their mental state. In this way, RMT could be used for real-time monitoring and detection of changes relevant to depression, contributing to more objective screening, improved symptom management, relapse-prevention, and delivery of just-in-time adaptive interventions. Initial work is promising, with evidence that RMT data features can distinguish those with depression from healthy controls and are associated with and predictive of standardised measures of depression symptom severity ^19–21^. However, depression itself, characterised by low motivation and energy, may make it more difficult to engage with RMT due to the requirement for daily interaction over long periods of time ^22,23^, and even less is known about the use of RMT for depression specifically in young people ^24^. Whilst narrative, systematic, and scoping reviews have previously been conducted ^25–29^, interest in the field, technological capabilities, and number of records available for inclusion increase steeply each year especially in the wake of the Covid-19 pandemic, with most digital mental health literature published between 2019 and 2022 (PubMed.gov). Despite this, regulation and standardisation remain inadequate, with few clinical trials on effectiveness or unintended outcomes and low continued engagement ^30–33^, as well as limited scrutiny of the realist and ethical considerations likely to significantly impact the benefits, implementation, and overall potential of RMT in the real-world. With pressure to find a scalable solution for the global mental health crisis and the movement further towards digital mental health, there is an ever-increasing danger that use of RMT may surge ahead of the evidence base ^34^. Therefore, the aim of this study was to conduct a realist review with lived experience involvement to determine the ways, for whom, in which contexts, and why RMT appear to work or not work for depression in young people aged 14 – 24 years and make recommendations for future research and practice.

## Methods

### Rationale for the Realist & Lived Experience Involvement Approach

Whilst traditional methods of review focus on determining the overall effectiveness of an intervention, a realist review seeks to determine the ways in which an intervention does or doesn’t work, when used by different individuals, in different contexts, and why, then make recommendations for how the intervention can be implemented and used most effectively ^35^. It is an iterative process encouraging consultation with key stakeholders throughout ^35^, who by virtue of their lived experience bring a wealth of additional expertise, knowledge, and insights important for real-world impact ^36^. In the present study, the realist review was conducted in in collaboration with two young, lived experience co-researchers from The McPin Foundation Young People’s Network (YPN) and in accordance with the Realist and Meta-narrative Evidence Syntheses: Evolving Standards (RAMESES) for quality and publication ^37^ Consultations were also held with the wider YPN to clarify scope, aid evidence synthesis, and develop the recommendations.

### Clarifying Scope & Search Processes

Through exploratory searches using broad search terms and selection criteria, snowball sampling, discussion with the YPN and co-researchers, and progressive focusing of scope, a final list of relevant intervention theories was produced. These were then categorised to form a theoretically based evaluative framework to be populated with evidence. The framework also formed the basis of the search strategy and bespoke data extraction template, with inputs from the co-researchers on sources of literature, search terms, refinement of selection criteria and data to be extracted. Searches were conducted across 10 electronic databases, including those with a more specialist focus on health technology (PubMed, Ovid – EMBASE, MEDLINE, PsychINFO and Global Health, Web of Science, Cochrane Library, IEEE Xplore, HTA database, ACM digital library) and 7 sources of grey literature (CADTH, NICE, WHO, ClinicalTrails.gov, ISRCTN registry, Gov.uk, rxiv.org), with a date restriction of Jan 1990 – current day (Aug 2021) reflecting the digital revolution. Search terms related to RMT, depression, young people, and the specific intervention theories included in the framework. Hand searches of reference lists of eligible literature and relevant systematic reviews were also conducted.

### Literature Selection Criteria & Appraisal

Following iterative searches and refinement, the final inclusion criteria were as follows: (i) peer-reviewed articles and grey literature; (ii) of any type including reviews, protocols, quantitative, qualitative and mixed-methods original research, and clinical trials displaying sufficient rigor and relevance; (iii) that assessed the use of RMT; (iv) for depression as the primary condition but other mental and physical comorbidities could be present; (v) specifically in young people aged 14 – 24 years old as defined by the World Health Organisation and United Nations ^38,39^, unless the sample was considering end-users of RMT involved in the care of young people (e.g., health care professionals, parents); (vi) from both clinical and community populations allowing inclusion of at-risk, sub-clinical young people who could deteriorate during the monitoring period. Literature where RMT was used solely to deliver an intervention without any remote measurement, focused on a specific type of depression (e.g., bipolar, perinatal, or postnatal), or did not include a standardized measure of depression (e.g., well-being scales) was excluded.

### Data Extraction & Evidence Synthesis

The bespoke data extraction template comprised a new row for each included literature record that may be relevant to several intervention theories, and columns for the input of extracted data relating to each intervention theory. Records were categorised and sorted by intervention theory then divided between the team lead (AW), co-researchers (GN, TS) based on their areas of interest, and two postdoctoral researchers (ZZ, MM) for data extraction. Initial evidence synthesis was undertaken by the team lead, with inferences then discussed with the YPN to ensure external validity against the lived experience of young people, and final review by the co-researchers to ensure perspectives were interpreted correctly without researcher bias. Where further testing and refinement of intervention theories was required, these literature search, selection, appraisal, extraction, and synthesis processes were repeated. In this way, the initial framework was gradually focused into a refined, realist intervention theory comprising multiple context-mechanism-outcome (CMO) configurations describing the ways, for whom, in which contexts, and why RMT appear to work or not work for depression in young people.

## Results

### Literature Inclusion & Characteristics

Of 6118 records identified, 104 were included in the final evidence synthesis (Fig. 1). Most investigated whether RMT data features could act as a proxy for depression symptom severity in young people (n = 36) and the acceptability and feasibility of doing so (n = 35). Data privacy ethics (n = 6) or possible unintended outcomes (n = 3) of real-time monitoring were severely under investigated given the growth of the field. Only 6 records originated from LMICs (Nepal, Columbia, Bangladesh, and Indonesia), and eligible grey literature was limited (n = 5), reflecting the current lack of regulation and government policy on the use of RMT for depression in young people.

**Fig. 1.**
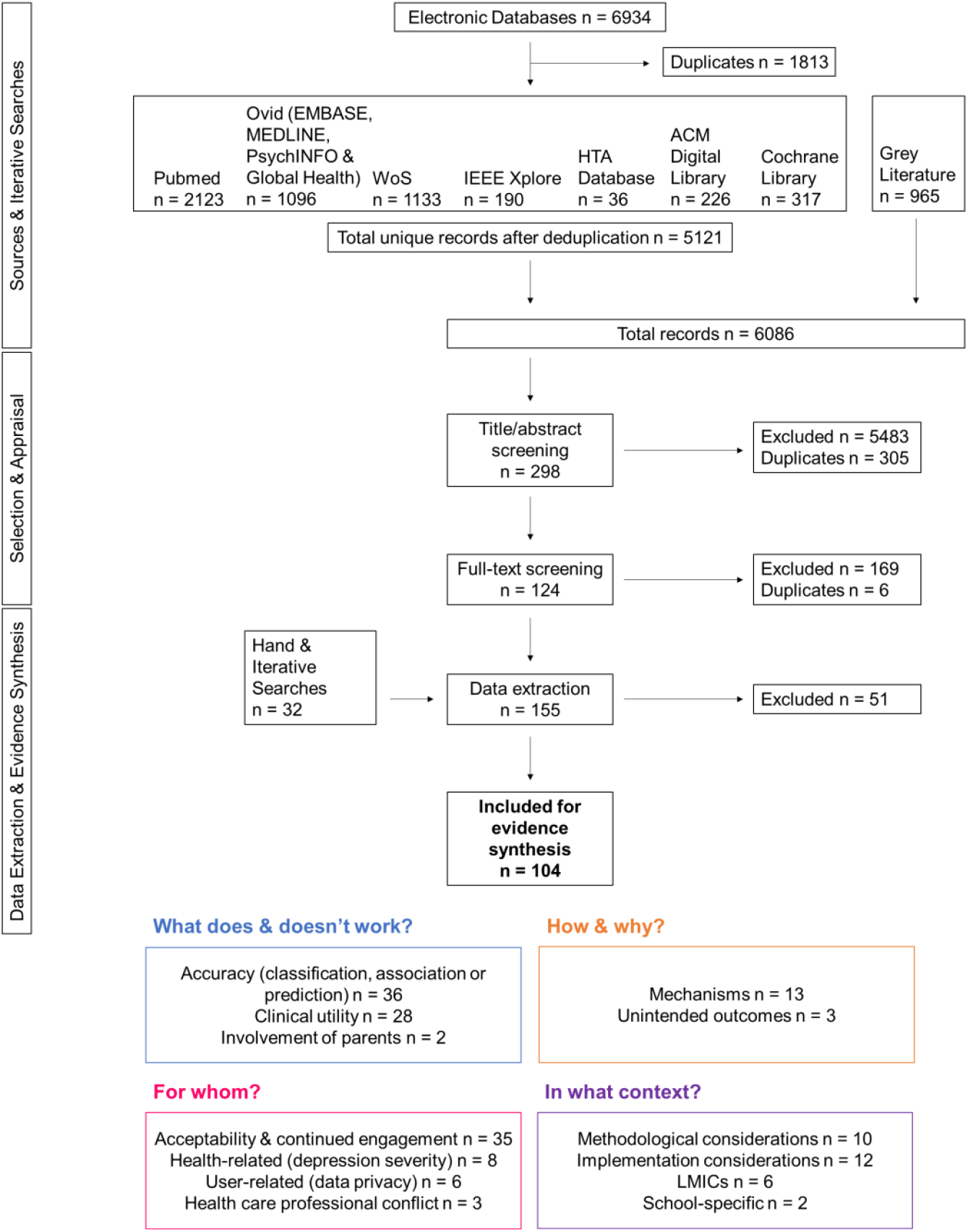
Flow diagram of searches, literature selection and grouping, data extraction and evidence synthesis.

### What does and does not work?

Out of the different types of RMT, smartphones and associated apps were the most prevalent in the literature (n = 52; internet-based platforms n = 23; wearables only n = 2; electronic diary n = 1; use in combination n = 22), as well as the most preferred by young people in the literature and during consultations, due to their ubiquity over other RMT ^29,40^. However, the choice of RMT ^41^ and type of smartphone operating system used (i.e., Android or iPhone) ^42^ were found to influence the specific data features identified, as well as overall accuracy as a proxy for depression symptom severity.

Regarding data collection, the continuous, unobtrusive nature of passive monitoring increased data quality and accuracy ^41^; however, this still required the young person to remember to keep the device charged and on their person every day for extended periods, with no technological issues (e.g., software crashes, GPS / Wi-Fi / Bluetooth dropout, drain on battery and data allowances) ^29,43^. As such, passive monitoring may be associated with overall lower adherence and greater decline in use over time compared with active monitoring ^43^.

Conversely, adherence to active monitoring required consideration of sampling frequency, but with conflicting evidence as to whether fewer or more EMA prompts was associated with greater adherence in young people with depression. Whilst a meta-analysis found an association between more prompts per day and higher adherence in clinical samples ^44^, prompt fatigue at the daily level was common across studies, with more robust engagement obtained in studies employing weekly or biweekly EMA protocols ^45,46^. It may be that the timing and content of prompts is more important than frequency, with the suggestion of smart prompts triggered when an individual is more likely to be receptive and able to respond ^47^, allowing convergence of prompt, motivation, and ability as the three elements required for a behaviour to occur ^48,49^. Provision of incentives, personalised feedback and gamification were also mentioned as ways to encourage continued engagement ^50^.

YPN members expressed that both types of data collection should be used in combination for a holistic approach, with passive monitoring to increase accuracy, active monitoring to provide context, and interactions kept brief but relevant / personalised and motivational in nature, and of an appropriate frequency and timing to balance accuracy and intrusiveness. Young people in the literature and during consultations also expressed increasing concern regarding the privacy, confidentiality, and ethical implications of the collection and sharing of digital data. Young people were much less likely to share digital data than psychosocial or biological data ^51^, with acceptability of certain digital data types particularly low e.g., audio, keystroke, social media, app usage and location data ^51,52^. They stressed the importance of transparent, informed consent and the need for user-control over what data is shared, how much, who with, and when ^53–55^. This would also allow the option to submit notes alongside for added context to avoid misinterpretation of the data or misrepresentation of the young person; however, one mentioned the risk of purposeful skewing of the data to reduce concern e.g., in comorbid eating disorder.

Once collected, RMT data features were able to distinguish those with depression from healthy controls and were associated with and predictive of standardised measures of depression symptom severity, with changes in sleep, mobility, smartphone use, social communication and EMA responses appearing to be the most promising for the detection of depression in young people.

### Sleep

A case-control study found young people with depression had significantly longer sleep latency, higher restlessness, shorter sleep duration and reduced sleep efficiency compared to with healthy controls, with a positive correlation between sleep latency and negative affect the following day ^56^. Conversely, remitted depressed had significantly longer sleep duration on work-free days compared with healthy controls ^57^ showing the importance of also collecting contextual data, but also that sleep disruption may be a persistent factor in depression that would be sensible to monitor in relapse-prevention. There was also conflicting evidence for the association between sleep duration and later depression symptom severity across longitudinal studies: one study found no significant correlation ^58^, one found a negative correlation ^59^; one found longer sleep duration at baseline but shorter sleep duration at follow-up associated with worse change in depression symptoms ^60^; one found longer sleep duration to predict greater anhedonia the following day, but when averaged across 2 weeks shorter sleep duration was predictive of greater anhedonia, reflecting the differential effects of acute versus chronic sleep deprivation in depression ^61^; however, for those still of school-age, longer sleep duration was associated with better sleep quality and predictive of lower depression symptom severity the following day, mediated by higher daytime energy level and moderated by parental enforcement of bedtimes and later school start times ^62,63^. Instead, greater variation in the sleep time series data was consistently found to be associated with greater depressive symptom severity [53–55]. This suggests that an overall irregular sleep pattern rather than any single discreet measure of sleep may be more indicative of worsening depression, especially in young people subject to changing demands and academic pressures ^60^.

### Mobility

Reduced mobility was also consistently found to be associated with greater depressive symptom severity in young people across a variety of different measures including step count ^64^, Bluetooth co-locations ^59^, GPS ^60^ and accelerometery ^58,66^. No measure was predictive of later depression symptom severity at 2-year follow-up; however, this was likely the result of using a 21-day snapshot of RMT data rather than continuous monitoring for the whole 2 years ^64^. Indeed, again, greater variation and lower predictability in daily activity duration time series data was associated with depression symptom severity only in those participating for more than 45 days, with functioning routines becoming less stable over time as depression worsens ^67^.

### Smartphone Use & Social Communication

Changes in patterns of smartphone use and social communication could be used to infer the reduced concentration, negative cognitive biases, and initial reassurance seeking ^68^ then subsequent social withdrawal observed in depression. Indeed, young people with depression were found to display significantly higher smartphone use specifically in study spaces ^58^, frequency of launch per app ^69^, use of social communication apps ^52,69^, and daily proportion of negative words typed within social communication apps ^70,71^ compared with healthy controls. Lower time spent in friends’ houses per visit ^66^ and reduced face-to-face conversation frequency and duration ^59^ were predictive of later depression symptom severity. Finally, as would be expected, EMA self-reported negative mood ^50,65^, as well as stress ^59,68^, self-esteem [61] and parent-reported negative mood ^73^ were found to be associated with and predictive of greater depression symptom severity.

RMT data features have been considered in isolation so far; however, their combination in linear regression and machine learning models increased predictive performance, with classification accuracy for standardised measures of depression ranging from 77 – 86% ^42,58,73–75^. Further improvements were obtained through contextual filtering of RMT data features (e.g., weekday / weekend sleep, daytime / evening conversation frequency) ^76^, normalisation relative to an individual’s own data ^77^, and inclusion of parent-report EMA responses ^47,73^.

### Clinical Utility

Whilst RMT data features certainly appear relevant to depression, there is less evidence for the clinical utility of this in improving depression outcomes in young people either through objective screening, symptom management, relapse-prevention, or just-in-time adaptive interventions. Of the 28 records pertaining to clinical utility only 10 were published studies, with the remaining 18 being registries and/or protocols of clinical trials yet to be completed ^78–96^.

Three studies investigated the use of RMT for one-off, opportunistic screening in young people. Comparison of RMT with face-to-face assessments of depression found fair to good concordance for the detection of positive screens of depression ^40,97–100^, as well as the detection of more psychosocial risk factors overall ^97,99^. This suggested increased disclosure of information without fear of judgement, which may aid quicker provision of more appropriate treatment.

The use of RMT for symptom management through self-monitoring and feedback to health care professionals was often rated as useful by young people ^101,102^ and resulted in some reduction of depression symptom severity ^103,104^. However, this reduction was not significantly different between self-monitoring and comparison groups ^46,103–111^, unless outliers were removed ^46^, or history of emotional abuse was considered ^112^. As such, RMT could aid symptom management in less severe cases but may not be sufficient for those with higher depression symptom severity and/or adverse childhood experiences ^46^.

Many highlighted the promise of RMT for relapse-prevention through detection of a “relapse-signature” and direct linkage to primary or secondary alert-based systems. RMT could even inform and directly deliver personalised digital micro-interventions that are highly focused towards the detected change requiring lower effort for purposeful engagement ^113^, or just-in-time adaptative interventions that adapt the provision of support to an individual’s changing state over time, such that the right type of support is delivered, at the right time, just when the individual needs it most ^74,75,114^. This would require understanding of which RMT data features precede relapse, their specificity for depression, their sensitivity in those already depressed, and the impact of detection on time to treat and depression outcomes, for which literature was limited, at least in young people. However, there were some small proof of concept studies for the prediction of day-to-day ^75^, or even hour-to-hour ^74^ fluctuations in depression symptom severity in clinical samples, leading to quick modification of care for the two patients detected and alerted as at risk ^45^, as well as several clinical trials underway^86,87,93,94^.

### How and why?

Whilst the overall impact of RMT on depression outcomes in young people remained inconclusive, self-monitoring specifically was consistently found to result in a greater increase in emotional self-awareness relative to comparison groups ^7,102,104,106,107,115^. Emotional self-awareness describes the ability to understand one’s own emotions and is often low is those with depression ^116^; however, it can be increased through improved understanding of the links between certain events or thoughts and later emotions, behaviours, and coping strategies. Indeed, this is a key component of cognitive behavioural therapy and forms the basis of many smartphone apps with EMA prompt protocols ^103,115,116^. Increased emotional self-awareness can lead to improvements in other important emotional processes, such as emotional granularity, controllability, and self-regulation ^117,118^, that may contribute to long-term behaviour change, more appropriate coping strategies and subsequent improvement in depression outcomes. Contrary to the concerns of healthcare professionals ^119^, self-monitoring was also found to increase readiness for and occurrence of help-seeking behaviours. In boys specifically, RMT were found to be an alternative outlet for expressing emotion, acting to reduce the stigma they felt they would receive if they talked about their emotions until they were ready to seek help ^105^.

Where there was feedback to the health care professional, RMT facilitated quicker and more efficient communication ^120^, prompted recall or the start of a conversations ^115^ with greater disclosure of information ^98^, increased understanding of the young person ^108^, allowed shared decision making, goal setting and tracking ^7,45,101,118,121^, and led to an overall increase in therapeutic engagement, alliance, and adherence ^80,115^. Finally, although limited, there was some evidence to suggest that use of RMT for depression in young people may have some unintended outcomes, such as worry in the absence of safeguarding when viewing pertinent data or occurrence of false negatives and/or positives ^40,52,122^; overreliance and overcontrol, particularly in those with co-morbid health anxiety, eating disorder or obsessive-compulsive disorder where self-monitoring could cause significant harm ^123^; burden of the data-driven nature, goal setting and behaviour change, and a sense of failure when not achieved ^124^, all of which could exacerbate depression symptoms. As such, through consideration of the underlying mechanisms of what does and does not work, RMT may be better used alongside current care pathways or to monitor and/or augment the effects of other interventions for depression in young people, rather than as an intervention itself. However, the question remains as to why adherence, accuracy, predictive performance and/or effectiveness may vary across young people and different contexts.

### For whom?

Throughout the literature, there was the assumption that RMT may be a natural fit for young people who have grown up during the digital revolution. Internet access and smartphone ownership in young people were relatively high and perspectives on the use of RMT for depression were generally positive ^43,52,101,102,106,110,120,122,125–128^. However, usage statistics were quite low ^126,129,130^, particularly for boys ^126,130^ despite the benefit they may gain from RMT through reduced perceived stigma and subsequent help-seeking behaviours ^105^. YPN members indicated that the low motivation and energy experienced during depression but requirement for daily interaction over extended periods of time played a significant part in their decision not to use RMT ^22,23,131^. Whilst no study specifically set out to investigate the impact of depression itself on the use of RMT and adherence rates as their main aim, it was still considered by some in related analyses, but with conflicting results. Five studies reported no significant effect of depression symptom severity on drop out [95,101], EMA prompt response rates ^44^, amount of entries / missing data ^67^ or exclusion due to missing data ^76^. Two studies suggested that those with greater depression symptom severity may be less likely to drop out ^47^, with continued engagement beyond the minimum time required by the study ^115^. Only 1 study found greater depression symptom severity, as well as treatment intensity, to be related to lower use of their smartphone app ^101^. Even though the lowest adherence rates reported were comparable to those of other interventions ^46,110,115^, it is important to note the impact of the heterogeneous, artificially inflated, or unreported quantitative measures of feasibility on the validity of adherence rates across the literature. With evidence that the level of adherence may be predictive of mental health outcomes at least in adults ^132^, there is a crucial need for standardisation to determine whether young people are likely to use RMT for depression, whether this translates to real-world settings outside of the study contexts and further investigate reasons why it may vary across individuals.

### In which contexts?

RMT were also assumed to be an increasingly ubiquitous and accessible resource that could be the much-needed scalable solution to the global mental health crisis. Whilst there was evidence that digital divides have declined in recent years, they were still pertinent, particularly in rural areas, LMICs and/or areas with higher accessibility but low digital literacy ^133,134^. As such, the widespread adoption of RMT could further perpetuate mental health inequalities and outcomes ^135^. Additionally, with high initial costs in the development and implementation of RMT, LMICs sometimes resort to making use of previously established RMT that may not be suitable for the context ^40^. Six studies investigated the use of RMT for depression in young people in LMICs, with a focus on adaptability to context, language, and culture key to their success ^40,100,134,136–138^. The factors found to influence the type of data that should be collected, and outcome measures included to make RMT more culturally compelling and accurate in local contexts were consideration of the role of families, locally defined experience of depression, and social determinants of mental health ^40,137,139,140^.

There were attempts to assess the feasibility of integrating RMT into primary and secondary mental health care services; however, differing digital infrastructures and capacities made the process complicated ^141,142^, uptake by consenting healthcare professionals was low (35/103, 34%) due to the changes in roles, responsibilities and training required ^107,115,143^, and there was a need to balance the expectations of young people, healthcare professionals and researchers regarding alert-based systems and capacity to respond ^7,40,101,111,115,119,122,128,143^. Whilst RMT could still be used to enhance the therapeutic relationship, its intended use in the early detection of deterioration for relapse-prevention is unfortunately highly unlikely to result in quicker time to treat under current resourcing and capacity levels.

Finally, the young lived experience co-researchers highlighted the potential challenges associated with real-time monitoring in schools, where young people may not always be able to keep RMT on their person since the use of smartphones and other technologies is often not allowed during class. There were two studies investigating the use of RMT in schools ^144,145^, with only 1 including school personnel ^144^, whose positive perspectives were likely the result of participation bias. Some studies were able to adapt data collection methods for the school context, such as fixed-time sampling with EMA prompts only occurring outside of school time; however, data was still missed that may be crucial to understanding the role of academic stressors in depression. Further investigation of acceptability and feasibility in a greater number of schools across different regions is required, with potential to integrate RMT alert-based systems into schools instead where there is somewhat more of a capacity for early detection and relapse-prevention ^145^. Overall, there is a need for increased implementation science to determine whether the significant challenges above can be overcome for the sustainable integration and scaling of RMT across contexts ^146^.

### Realist Intervention Theory of RMT for Depression in Young People

Evidence synthesis allowed gradual refinement of the initial framework into a realist intervention theory of the ways, for whom, in which contexts, how and why RMT appear to work or not work for depression in young people aged 14 – 24 years, as shown in Fig.2

**Fig. 2.**
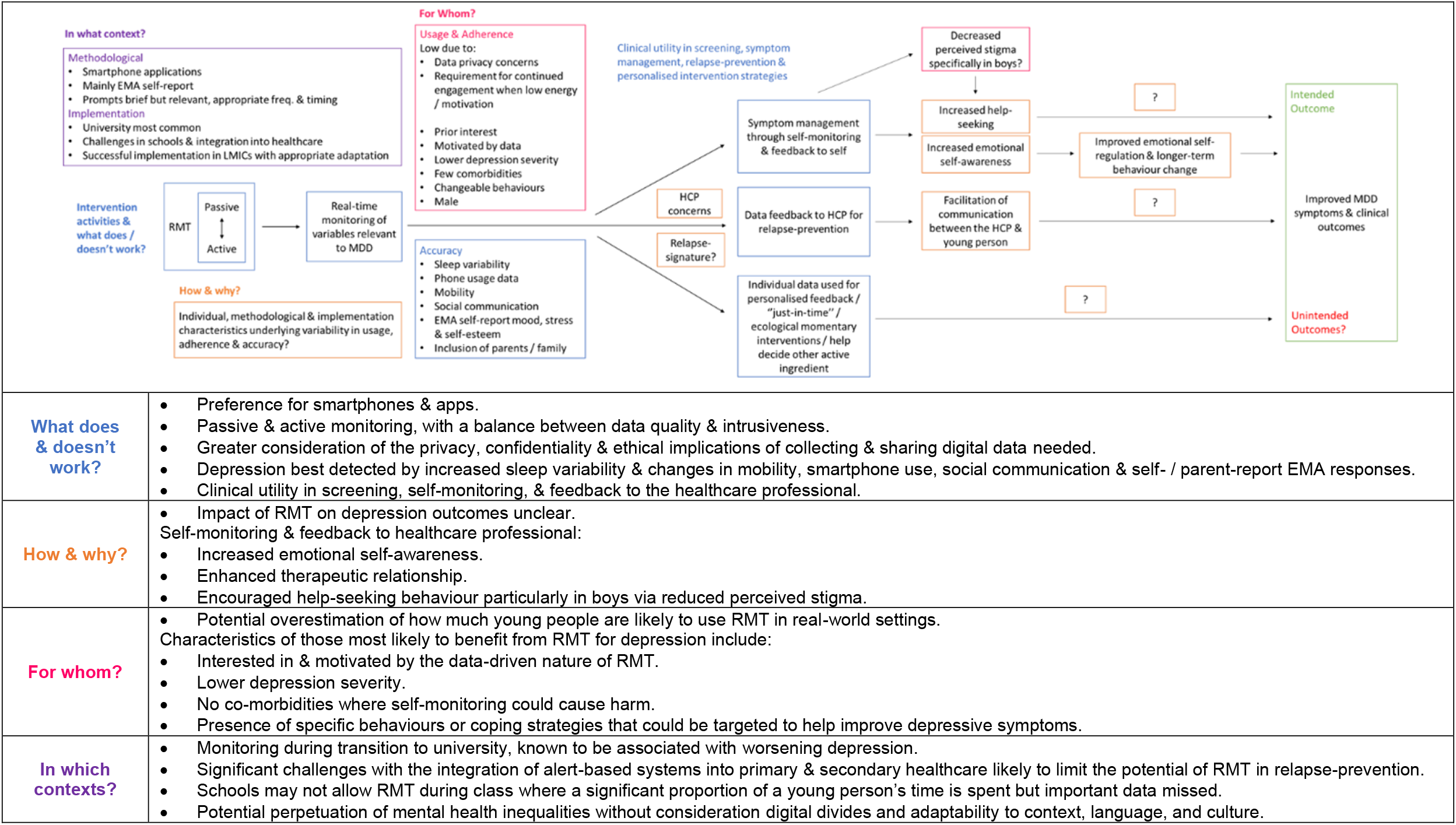
Realist intervention theory of RMT for depression in young people and explanation of the patterns observed. Question marks indicate areas where evidence is lacking and there is a need for further research.

## Discussion & Recommendations

There has been a significant movement towards digital mental health in recent years, with the potential for RMT to be the scalable solution to the global mental health crisis ^34^. However, in the wake of this steep growth, regulation and standardisation remain inadequate, with limited understanding of the effect on depression outcomes nor scrutiny of the realist and ethical considerations likely to significantly impact the benefits, implementation, and overall potential of RMT in the real-world ^30–33^. Therefore, a realist review was conducted in collaboration with The McPin Foundation YPN through which a refined, realist intervention theory was successfully produced explaining the ways, for whom, in which contexts, how and why RMT appear to work or not work for depression in young people aged 14 – 24 years.

It also highlighted gaps where very limited evidence was available, as well as several common issues that appeared across much of the literature reviewed including: convenience sampling and selection bias with those young people more interested in RMT more likely to take part; reliance on self-report in community samples with few reporting the number above / below clinical cut-offs; potentially inappropriate statistical analyses for the low sample sizes but large time series data sets ^147^; short study duration (7 days to 12 months at most) or snapshot monitoring periods (2 – 3 weeks) in multiyear studies despite estimated relapse rates to be only 5% within the first 6 months, 12% by 12 months, 40% by 2 years, then 70% by 5 years ^148–150^; low number of predictor variables and little consideration of the many factors that could have confounding, mediating or moderating effects over the length of the monitoring period; potentially conflicting indicators of depression symptom severity due to the diversity of experience overlooked ^76,77,122^; heterogeneity, artificial inflation, or non-report of quantitative measures of feasibility making comparison across studies difficult, especially with regards to adherence ^27,44^; limited investigation of specificity, sensitivity, and impact on time to treat and depression outcomes; and overall little attempt to assess and explain variability in adherence, accuracy and predictive performance across individuals ^74^. With the evidence base as it stands and important insights from those with lived experience, we make the following recommendations for the use of RMT for depression in young people:

### Recommendations for future research

Before moving forward, ethical procedures for the collection, sharing, and use of digital data need to be reviewed and updated, quantitative measures of feasibility and depression outcomes need to be standardised, and other methodological issues need to be rectified to increase the validity of results. Future research should then focus on (i) potential unintended outcomes of real-time monitoring; (ii) predictive investigations to determine which, if any, RMT data features precede and have sensitivity and specificity for depression relapse; (iii) use of RMT to inform and deliver digital micro-interventions and just-in-time adaptive interventions; (iv) impact of the use of RMT on time to treat and depression outcomes in young people; and (v) implementation science to determine whether the significant challenges can be overcome.

### Recommendations for practice

Without the above in place, the current best use of RMT for depression in young people is not as an intervention itself but as part of a blended, stepped-care approach, with use throughout the care pathway from opportunistic screening, watchful waiting, triage, and placement on waiting lists, to during treatment to monitor effectiveness, increase emotional self-awareness and enhance the therapeutic relationship.

## Data Availability

All data produced in the present work are contained in the manuscript.

## Acknowledgements

This work was funded by a Wellcome Trust Mental Health ‘Active Ingredients’ commission awarded to AW at KCL. We would like to thank The McPin Foundation and their Young People’s Network for their significant contributions to the project, manuscript, and dissemination. VM and AW are also supported by a MQ Brighter Futures grant [MQBF/1 IDEA], and VM by the National Institute for Health Research Biomedical Research Centre at South London and Maudsley NHS Foundation Trust and King’s College London (NIHR Maudsley BRC). The views expressed are those of the authors and not necessarily those of the Wellcome Trust, MQ, NHS, NIHR, Department of Health and Social Care, or King’s College London.

## Abbreviations

EMA: ecological momentary assessment
LMICs: low-to-middle income countries
RMT: remote measurement technologies
YPN: Young People’s Network

